# Health Belief Model and Experiential Avoidance in Relation to PTSD Symptoms Among Healthcare Workers in Ekiti State, Nigeria: A Structural Model Analysis

**DOI:** 10.64898/2026.05.08.26352756

**Authors:** Dogbahgen Alphonso Yarseah, Olu Francis Ibimiluyi, Falana Bernard Akinlabi, Ayodele Chirstian, Bello Folaranmi Fatai, Ololade Omolayo Ogunsanmi, Jedege Oluwadamilola

**Affiliations:** Ekiti State University, Faculty of Education Department of Guidance and Counseling Ekiti State, Ado Ekiti, Nigeria, Country: Liberia; Ekiti State University, Faculty of Education, Department of Guidance and Counseling Ekiti State, Ado Ekiti, Nigeria; Cavendish University of Uganda Department of International Relations Kampala, Uganda; Department of Public Health Babcock University, Ogun State, Ilishan-Remo, Nigeria

**Keywords:** Health Belief Model, Experiential Avoidance, PTSD, Healthcare Workers, PLS-SEM, Mediation, Moderation

## Abstract

**Background:** Healthcare workers are at increased risk of post-traumatic stress disorder (PTSD) due to prolonged exposure to high-stress clinical environments. Although the Health Belief Model (HBM) has been widely used to explain health behaviors, its application to psychological outcomes such as PTSD remains limited. The role of cognitive-emotional processes, particularly experiential avoidance, in linking health beliefs to trauma symptoms is not well understood.

**Methods:** This study adopted a quantitative cross-sectional design to collect data from 475 healthcare workers in Ekiti State, Nigeria. Participants completed standardized measures assessing Health Belief Model constructs, experiential avoidance, and PTSD symptoms. Data were analyzed using Partial Least Squares Structural Equation Modeling (PLS-SEM), with bootstrapping used to test direct, indirect (mediation), and moderation effects. Cluster analysis was also conducted using SPSS to validate differences in PTSD symptom severity across psychological constructs and demographic variables.

**Results:** Experiential avoidance significantly predicted PTSD symptoms (β = 0.395, 95% CI [0.231, 0.565]). HBM constructs were negatively associated with experiential avoidance (β = - 0.198, 95% CI [-0.270, -0.108]) and PTSD symptoms (β = -0.119, 95% CI [-0.216, -0.006]). Mediation analysis indicated that experiential avoidance partially mediated the relationship between HBM constructs and PTSD (β = -0.078, 95% CI [-0.132, -0.037]), with a total effect of - 0.197. Age moderated the relationship between HBM and experiential avoidance (β = -0.114, 95% CI [-0.207, -0.025]) as well as the indirect pathway to PTSD. Sex significantly predicted PTSD symptoms (β = 0.358, 95% CI [0.214, 0.501]). Cluster analysis showed that experiential avoidance and perceived barriers significantly differentiated high and low PTSD symptom groups.

**Conclusion:** The findings support a conditional cognitive-emotional model in which Health Belief Model constructs influence PTSD symptoms both directly and indirectly through experiential avoidance. Demographic factors shape the strength of these relationships, while perceived barriers and experiential avoidance emerge as key determinants of trauma-related distress among healthcare workers.

## INTRODUCTION

The world was gripped with fear following the emergence of COVID-19 in Wuhan, Hubei Province, China on 31 December 2019, which subsequently led to the declaration of a pandemic by the World Health Organization (WHO) on 13 January 2020 [1]. The COVID-19 pandemic resulted in substantial global morbidity and mortality, with millions of confirmed cases and significant fatalities reported worldwide [1]. In Africa, the virus was first confirmed in Egypt on 14 February 2020 and rapidly spread to Nigeria within a week, where thousands of cases and several hundred deaths were recorded [2–3].

The rapid emergence of the pandemic created widespread fear, uncertainty, and psychological distress within the Nigerian population. These effects were intensified by structural and infrastructural challenges, including limited access to electricity, television, and radio in some regions, which constrained timely access to accurate health information. Government-imposed containment measures, including lockdowns and closure of public institutions, further disrupted daily life and contributed to increased psychological distress, hopelessness, and anxiety among the general population.

Among the most affected groups were healthcare workers, who were exposed to heightened occupational demands within an already fragile healthcare system. Nigeria’s healthcare system is characterized by limited resources, weak referral structures, and low health insurance coverage, all of which increase occupational stress during public health emergencies [4]. In addition, the healthcare workforce remains critically inadequate, with an estimated doctor-to-patient ratio of approximately 1:2753, significantly limiting service delivery capacity during crises [5–6]. These conditions place healthcare workers at elevated risk of psychological strain, particularly during pandemics.

Emerging evidence suggests that healthcare workers exposed to traumatic clinical environments are at increased risk of developing post-traumatic stress disorder (PTSD), characterized by symptoms such as re-experiencing, avoidance, and hyperarousal [7–8]. PTSD is not only a consequence of trauma exposure but is also shaped by cognitive appraisals and emotional regulation processes [9]

The Health Belief Model (HBM) [10–11] provides a cognitive framework for understanding how individuals perceive and respond to health threats. It posits that perceptions of susceptibility, severity, and barriers influence psychological and behavioral responses. In the context of healthcare workers during COVID-19, these beliefs may shape how individuals interpret risk exposure and manage stress-related experiences.

However, cognitive appraisal alone may not fully explain the development of PTSD symptoms. Experiential Avoidance (EA), defined as the tendency to avoid or suppress unwanted internal experiences, has been identified as a key maladaptive coping mechanism within psychological flexibility theory [9]. High levels of experiential avoidance may prevent effective emotional processing of traumatic experiences, thereby increasing vulnerability to PTSD symptoms.

Integrating these frameworks, this study proposes that health beliefs influence PTSD symptoms both directly and indirectly through experiential avoidance. Specifically, maladaptive coping processes may mediate the relationship between cognitive appraisals of health threats and psychological outcomes.

Furthermore, individual differences may shape these relationships. Age may influence the strength of associations between health beliefs and experiential avoidance due to developmental differences in cognitive appraisal, coping flexibility, and emotional regulation across the lifespan. Similarly, sex differences in emotional processing and stress reactivity suggest that the relationship between experiential avoidance and PTSD symptoms may vary between males and females.

### Posttraumatic stress disorder and experiential avoidance

These structural and systemic deficiencies placed healthcare workers at increased risk of posttraumatic stress disorder (PTSD) such as re-experiencing, hypervigilence, and avoidance. Studies in Asia, Europe and some countries in Africa have demonstrated HCWs suffered from severe PTSD during the COVID-19 pandemic (7-8,12). Evidence from a meta-analysis indicates that approximately 13.52% of frontline healthcare workers experienced PTSD during the COVID-19 pandemic (95% CI: 9.06–17.98), reflecting a considerable psychological burden associated with prolonged exposure to pandemic-related stressors, however, to our knowledge research on PTSD among healthcare workers in Nigeria most especially Ekiti State is not known.

This high prevalence suggests that healthcare workers are vulnerable to trauma-related outcomes, potentially influenced by maladaptive coping mechanisms such as experiential avoidance and cognitive appraisals related to health threats [13]. In addition, North et al [14] to provides an in-depth nosological consideration of the diagnosis of PTSD showed that the prevalence of PTSD ranges from 12% to 96% in hospitalized COVID-19 patients, 4% to 73% in healthcare workers and 3% to 67% of general populations. He and colleages later argued that diagnostic frameworks such as ICD-11 provide broad criteria for trauma exposure, some scholars caution against overextending the definition of trauma in the context of COVID-19. For instance, North et al [14] argue that only individuals directly exposed to life-threatening events—such as severe illness, intensive care conditions, or witnessing death—should be considered at risk of developing PTSD. This distinction is important to avoid conflating general psychological distress with trauma-related psychopathology.

In this context, healthcare workers represent a uniquely vulnerable population, as they are frequently exposed to critical and life-threatening situations during the COVID-19 pandemic. Their frontline role involves direct interaction with severely ill patients, repeated exposure to mortality, and high-risk clinical environments. Consequently, focusing on healthcare workers allows for a more accurate examination of PTSD within a population that meets the clinical threshold for trauma exposure.

Beyond nosological considerations that emphasize trauma exposure as a precursor to post-traumatic stress disorder (PTSD), cognitive and information-processing mechanisms also play a critical role in the development of PTSD. One such mechanism is experiential avoidance (EA), which reflects maladaptive responses to internal psychological experiences[9].

Experiential avoidance refers to the tendency to avoid or suppress unwanted internal experiences such as thoughts, emotions, memories, and physical sensations[15]. It is characterized by an unwillingness to remain in contact with these internal experiences, leading individuals to engage in cognitive and emotional strategies aimed at controlling or escaping them[16].

Empirical evidence strongly supports the role of experiential avoidance in PTSD. For instance, Yarseah et al. [17] reported that experiential avoidance was a strong predictor of PTSD symptoms (β = 0.814), explaining 67% of the variance (R² = 0.67). Similarly, experiential avoidance has consistently been identified as a maladaptive coping strategy that contributes to the development and maintenance of PTSD symptoms [14–17].

Mechanistically, experiential avoidance has been shown to heighten sensitivity to situational cues, increasing susceptibility to anxiety and panic reactions [18]. It may also divert attention away from task-relevant activities [19], bias cognitive evaluation through memory distortions, and intensify emotional responses while reducing pain tolerance [20–21]. These processes interfere with adaptive emotional regulation and increase vulnerability to trauma-related psychopathology.

Given these mechanisms, experiential avoidance can be conceptualized as a key psychological process involved in the development of PTSD. Specifically, it represents a pathway through which external stressors are translated into internal psychological distress. By disrupting emotional processing and promoting maladaptive cognitive responses, experiential avoidance may explain how exposure-related factors ultimately lead to PTSD symptoms among healthcare workers.

### Health Belief Model and PTSD

On the other hand, the relationship between health belief model and PTSD remains limited with few studies using additive effect [17]. Health-related behaviors are shaped by both overt and covert processes. Overt behaviors refer to observable actions such as adherence to protective measures, whereas covert processes involve internal cognitive and emotional responses, including perceptions of risk, fear, and appraisal of health threats [23]. These internal processes play a critical role in how individuals interpret and respond to health-related situations. In behavioural terms, overt behaviours are directly observable, whereas covert behaviours occur internally and include private events such as thinking and imagining Within this context, the Health Belief Model provides a useful framework for understanding how such covert processes influence health behavior. The model posits that individuals’ actions are guided by their perceptions of susceptibility, severity, benefits, and barriers, which are fundamentally cognitive evaluations of health threats [11]. Thus, health beliefs can be understood as structured representations of covert processes that shape behavioral responses to health risks.

Previous research has demonstrated a significant association between the Health Belief Model (HBM) and post-traumatic stress disorder (PTSD), particularly among medical students, where stronger health beliefs were associated with lower PTSD symptoms[24–25,17]. However, these studies have primarily focused on direct relationships between cognitive health perceptions and PTSD outcomes, without examining the underlying psychological mechanisms that explain this association.

In addition, empirical evidence on the HBM–PTSD relationship remains limited, particularly in high-risk occupational groups such as healthcare workers like Ekiti Stae. More importantly, little is known about the processes through which health beliefs translate into psychological distress. To address this gap, the present study extends existing literature by introducing experiential avoidance as a potential mediating mechanism through which health beliefs influence PTSD among healthcare workers. This approach provides a more comprehensive explanation of how cognitive health perceptions are translated into maladaptive psychological outcomes.

### Experiential Avoidance and Health belief model

The Health Belief Model (HBM) and Experiential Avoidance (EA) may be conceptually related despite representing distinct psychological processes [26–27]. While HBM reflects individuals’ cognitive appraisals of health threats and behaviors, EA is a verbally mediated process involving an unwillingness to remain in contact with negatively evaluated private experiences—such as thoughts, memories, emotions, and bodily sensations—and deliberate efforts to control their form, frequency, or intensity, even when doing so leads to actions that are incongruent with valued goals [28–29]. EA often functions as a short-term coping mechanism that provides immediate emotional relief; however, over time, its use becomes negatively reinforced, increasing the likelihood of repeated reliance on avoidance strategies when facing similar situations [15]. Research has shown that individuals who frequently engage in experiential avoidance may experience a progressive disengagement from valued life goals and a persistent psychological struggle [30,31].

Within this framework, constructs of the Health Belief Model particularly perceived severity and perceived susceptibility (often referred to as perceived seriousness) [32] may contribute to emotional and psychological burden among healthcare workers. Additionally, perceived barriers, which encompass obstacles encountered during health crises such as the COVID-19 pandemic, may further intensify this burden. The HBM has been widely used to explain individual perceptions and attitudes toward disease prevention and treatment adherence [32–34], positing that health-related behaviors are influenced by perceived severity, susceptibility, and barriers to action [35–37]. Since its development, the model has been expanded to include constructs such as perceived benefits, self-efficacy, and cues to action [338], and it has been extensively applied in preventive health contexts, including vaccination research [35].

Given that HBM constructs are inherently linked to perceptions of threat and risk, they may evoke emotional responses that increase psychological distress. Recent research applying the HBM to mental health discourse on social media indicates that higher perceived severity is associated with increased negative emotional expression [39]. Although this work does not directly examine experiential avoidance, heightened emotional distress has been theoretically associated with greater reliance on avoidance-based coping strategies. Furthermore, empirical evidence indicates that perceived susceptibility and perceived severity are strong predictors of perceived risk within the HBM framework [40]. While increased risk perception can promote adaptive health behaviors, in high-stress contexts it may also lead to maladaptive coping responses, such as experiential avoidance.

In this context, it is plausible that HBM constructs are associated with experiential avoidance, as individuals may attempt to avoid distressing health-related thoughts and emotions. Conversely, experiential avoidance may influence how individuals perceive health threats and barriers. Empirical evidence by Yarseah et al. [17] supports a significant relationship between HBM and EA; however, further studies are required to clarify the direction and magnitude of this association. Additionally, recent findings suggest that HBM constructs are emotionally laden and related to experiential processes [17], further supporting a potential link between HBM and EA.

### Mediating Role of Experiential Avoidance (EA) in the Relationship Between Health Belief Model (HBM) and PTSD

Building on the established relationship between Health Belief Model (HBM) constructs and experiential avoidance (EA), it is important to examine the mediating mechanism through which these variables influence psychological outcomes such as post-traumatic stress disorder (PTSD). Experiential avoidance is a psychologically inflexible coping process characterized by efforts to avoid or suppress distressing internal experiences, including thoughts, emotions, and memories [30–32].

Notably, avoidance is also a core component of PTSD. Individuals with PTSD often engage in avoidance of trauma-related thoughts, emotions, people, and situations. Within the framework of psychological flexibility theory, such avoidance behaviors have been conceptualized as manifestations of experiential avoidance [41–42]. This perspective suggests that psychological flexibility interferes with adaptive functioning and the pursuit of valued life goals, including maintaining meaningful social relationships [30–31].

Empirical evidence further indicates that experiential avoidance frequently mediates the relationships between emotional dysregulation, trauma exposure, and maladaptive behaviors, including substance use. Elevated levels of experiential avoidance have also been associated with impulsivity, psychiatric comorbidity, and poorer treatment adherence and outcomes [31]. Interventions targeting experiential avoidance, particularly Acceptance and Commitment Therapy (ACT), have demonstrated effectiveness in reducing avoidance patterns and improving psychological functioning. Collectively, these findings support the view that experiential avoidance operates as a transdiagnostic process underlying vulnerability to, and persistence of, a range of psychological disorders.

The mediation hypothesis posits that experiential avoidance explains the relationship between Health Belief Model (HBM) constructs and PTSD symptoms. Within the Health Belief Model, cognitive appraisals such as perceived susceptibility, severity, and barriers influence how individuals respond to perceived health-related threats [26–27]. These cognitive evaluations have been associated with psychological distress in high-stress occupational settings.

Experiential avoidance, as conceptualized in Acceptance and Commitment Therapy, refers to attempts to avoid or suppress unwanted internal experiences, which can temporarily reduce distress but contribute to long-term psychological dysfunction [31–32]. Empirical evidence shows that experiential avoidance is strongly linked to PTSD symptoms and other forms of trauma-related distress [17,41].

Although the full mediating pathway has not been extensively tested, prior literature supports the individual relationships between HBM constructs and distress, and between experiential avoidance and PTSD. Therefore, experiential avoidance is theoretically positioned as a mechanism through which cognitive health beliefs influence PTSD symptoms among healthcare workers in Ekiti State.

Age has been widely discussed in psychological and health behavior literature as a relevant demographic factor influencing stress responses, cognitive appraisal, and coping mechanisms. The Strength and Vulnerability Integration Theory [43] proposes that emotional regulation capacities tend to improve with age due to greater motivational control and accumulated life experience. Similarly, lifespan developmental theories suggest that older adults are more likely to adopt adaptive coping strategies and demonstrate greater emotional stability compared to younger individuals [44–45].

Within health behavior research, the Health Belief Model also suggests that cognitive appraisals such as perceived susceptibility, severity, and barriers may be interpreted differently across age groups, potentially influencing health-related decision-making and emotional responses [26–27]. Younger individuals are often reported to exhibit higher emotional reactivity and less stable coping strategies, whereas older adults tend to rely on more experience-based and emotion-regulation-focused coping mechanisms.

However, evidence across studies remains mixed, as age-related differences are often influenced by contextual stressors, particularly in high-demand environments where chronic stress exposure may reduce typical developmental advantages in emotional regulation.

### The moderating roles of agerange and Sex among HCWs

Healthcare workers (HCWs) experienced substantial psychological strain during the COVID-19 pandemic, with increased risks of occupational stress, emotional exhaustion, and trauma-related symptoms. High job demands and sustained patient interaction have been consistently linked to elevated psychological burden among health professionals [46]. Empirical evidence further demonstrates variability in stress levels across professional groups, with nurses and frontline healthcare staff reporting the highest levels of distress compared to other medical personnel [47].

Importantly, demographic differences in psychological outcomes have been widely documented. Female healthcare workers consistently report higher levels of psychological distress, anxiety, and PTSD-related symptoms compared to males [48–50]. In addition, age has been identified as a relevant factor, although findings remain mixed, with some studies suggesting greater vulnerability among younger workers, while others highlight increased strain among older or more experienced staff [51–52]. Occupational characteristics, including years of practice and professional role, have also been associated with variations in psychological responses, including experiential avoidance and health-related beliefs during the pandemic [17, 53–54].

However, despite this growing body of evidence, most studies have examined demographic variables such as age and sex as independent predictors of psychological outcomes rather than as factors that may influence the relationships among key psychological constructs. Given that Health Belief Model (HBM) constructs and experiential avoidance (EA) are central to understanding PTSD risk, it is plausible that demographic characteristics may shape how these variables interact. Specifically, age and sex may influence the strength or direction of the relationships between HBM constructs, experiential avoidance, and PTSD symptoms.

Therefore, the present study extends prior research by examining **the** moderating roles of age range and sex in the relationship between HBM constructs, experiential avoidance, and PTSD symptoms among healthcare workers, thereby addressing an important gap in the literature [52,55].

### Theoretical Framework

This study is grounded in the Health Belief Model (HBM), which proposes that individuals’ cognitive appraisals of health threats influence their emotional and behavioral responses. Within the context of trauma exposure, health-related beliefs may shape how individuals interpret and respond to distressing experiences, thereby influencing psychological outcomes.

Complementing this framework, Experiential Avoidance (EA) refers to the tendency to avoid or suppress unwanted internal experiences such as thoughts, emotions, or memories. High levels of experiential avoidance are considered maladaptive, as they may interfere with emotional processing of trauma and contribute to the persistence of Post-Traumatic Stress Disorder (PTSD) symptoms.

Based on this integrated perspective, PTSD is conceptualized as an outcome influenced by both cognitive (health beliefs) and emotional regulation (experiential avoidance) processes. Specifically, adaptive health beliefs are expected to reduce maladaptive avoidance tendencies, thereby lowering PTSD symptom severity, whereas higher experiential avoidance is expected to intensify PTSD symptoms.The model further incorporates demographic variables as boundary conditions. Age is conceptualized as a contextual factor that may influence how health beliefs translate into experiential avoidance, reflecting developmental differences in cognitive appraisal and coping flexibility. Sex is considered a potential determinant of PTSD symptom severity, given established evidence of gender differences in trauma exposure, emotional regulation, and stress reactivity. Overall, the study adopts an integrated cognitive–behavioral framework in which health beliefs influence PTSD outcomes both directly and indirectly through experiential avoidance, while demographic characteristics condition these relationships.

This study aims to examine the relationships among Health Belief Model (HBM) constructs, experiential avoidance, and PTSD symptoms. In particular, it evaluates the direct and indirect (mediation) pathways linking health beliefs to PTSD through experiential avoidance, and assesses whether these relationships are conditioned by demographic factors, specifically age and sex.

## Method

### 1. Ethical Considerations

Ethical approval for this study was obtained from the Human Research and Ethics Committee of the Federal Teaching Hospital, Ido-Ekiti (Approval No: ERC/2020/03/22/7965). Additional authorization was granted by the Research Training and Service Unit of the Federal Ministry of Health. All procedures involving human participants were conducted in accordance with relevant national guidelines and the Declaration of Helsinki.

### 2. Research Design

This study employed a quantitative cross-sectional design conducted among healthcare workers during the COVID-19 pandemic in 2020 in Ekiti State, Southwest Nigeria. The design was appropriate for examining associations between health beliefs, experiential avoidance, and PTSD symptoms at a single point in time.

### 3. Setting and Participants

The study was conducted in Ekiti State, Southwest Nigeria, a region that experienced a high burden of COVID-19 infections during the early phase of the pandemic. Ekiti State has a population of approximately 5.4 million and is predominantly inhabited by the Yoruba ethnic group with a well-established healthcare system comprising primary, secondary, and tertiary healthcare facilities across both public and private sectors.

Ekiti State has 16 Local Government Areas. Of these two urban Local Government Areas (LGAs), Ado and Ido-Osi, were purposively selected for this study because they host the state’s major tertiary healthcare facilities, which played a central role in the management of COVID-19 cases during the pandemic. These areas were considered critical settings due to their high patient load and increased exposure risk among healthcare workers. Participants comprised healthcare workers, including medical doctors, nurses, and other frontline professionals working in healthcare facilities within the selected LGAs. These individuals were directly involved in patient care during the COVID-19 pandemic and were therefore at heightened risk of psychological distress, including PTSD symptoms.

### 4. Participants and eligibility criteria

A total of 475 healthcare workers (HCWs) were recruited from healthcare facilities in Ado-Ekiti and Ido-Osi Local Government Areas of Ekiti State, Southwest Nigeria. The sample comprised medical doctors, nurses, pharmacists, and other allied health professionals involved in healthcare service delivery during the COVID-19 pandemic. Participants were recruited using a snowball sampling technique due to restricted access to healthcare facilities and the high workload of frontline workers during the pandemic. Initial participants were identified through professional networks and were asked to refer eligible colleagues.

Eligible participants were HCWs aged 18 years or older who were actively employed in Ado-Ekiti or Ido-Osi and involved in direct or indirect patient care during the COVID-19 pandemic. HCWs were excluded if they were employed in any of the other 14 Local Government Areas of Ekiti State or were not actively engaged in healthcare service delivery during the study period. HCWs who provided incomplete responses were also excluded from the study.

#### Data Collection Procedure

Data were collected between May 5 and June 18, 2020, during the early phase of the COVID-19 pandemic. Two urban Local Government Areas (LGAs), Ado and Ido-Osi in Ekiti State, Southwest Nigeria, were purposively selected for this study due to their high concentration of healthcare facilities, including the state’s tertiary hospitals that were actively involved in the management of COVID-19 cases.

Healthcare workers were recruited using a snowball sampling technique to facilitate access to frontline professionals under the restricted and high-risk conditions of the pandemic. Initial participants were identified through professional networks within selected healthcare facilities and were subsequently asked to refer eligible colleagues.

Data were collected using a structured self-administered questionnaire distributed through both paper-based and limited online formats, depending on accessibility. Given the contagious nature of COVID-19, strict safety protocols were observed during data collection, including the use of personal protective equipment (PPE), adherence to physical distancing guidelines, and minimization of direct contact.

Participation was voluntary, and informed consent was obtained from all participants prior to data collection. A total of 475 completed responses were obtained and included in the final analysis.

## Measures

### PTSD Symptoms

PTSD symptoms were assessed using the PTSD Checklist – Civilian Version (PCL-C), a widely used self-report measure developed by Weathers et al [56], developed based on DSM-IV criteria and comprising re-experiencing, avoidance, and hyperarousal symptom clusters. Although the PTSD Checklist used in this study is based on DSM-IV criteria, evidence from psychometric and crosswalk studies indicates substantial comparability between DSM-IV and DSM-5 PTSD measures. Empirical work has demonstrated strong overlap in symptom measurement and scoring equivalence across versions, with high correspondence observed in both clinical and non-clinical samples [57–58]. This suggests that DSM-IV–based PTSD symptom measures remain suitable for assessing overall PTSD symptom severity in research contexts, particularly in occupational and non-diagnostic populations. [57]. This supports the continued use of DSM-IV-based instruments in contemporary epidemiological and comparative research. In this study, PTSD was modeled as a reflective latent construct and demonstrated strong internal consistency (α = 0.875; CR = 0.923) and convergent validity (AVE = 0.800). Discriminant validity was also established, with HTMT values below the recommended threshold of 0.85 (maximum = 0.508).

### Experiential Avoidance

Experiential avoidance was measured using the Acceptance and Action Questionnaire-II (AAQ-II) developed by Bond et al.[41]. The AAQ-II is a 7-item scale assessing psychological inflexibility and avoidance of unwanted internal experiences using a Likert-type response format. High scores on the AAQ-2 are reflective of greater experiential avoidance and immobility, while low scores reflect greater acceptance and action. Test scores on the AAQ-II have demonstrated good internal consistency and test-retest reliability in community samples [41]. Prior research has demonstrated strong psychometric properties, including good internal consistency (α ≈ 0.84), test–retest reliability (≈ 0.81) [41], and strong convergent validity with measures of psychological distress. he AAQ-II has demonstrated strong psychometric properties in prior research, including good internal consistency and test–retest reliability [41]. In the present study, the construct demonstrated strong internal consistency (α = 0.852; CR = 0.888) and adequate convergent validity (AVE = 0.532).

### Health Belief Model (HBM)

The Health Belief Model (HBM) is a theoretical framework used to explain health-related behavior (Rosenstock [27]; Becker [59]). In this study, it was operationalized through three core constructs: perceived susceptibility, perceived severity, and perceived barriers. As HBM is a conceptual model rather than a standardized scale, its measurement depends on contextual operationalization. However, previous studies have reported acceptable internal consistency for its components when properly adapted, typically exceeding Cronbach’s alpha values of 0.70[60]. For this recent study the internal consistency (α = 0.717; CR = 0.900 ) and convergent validity (AVE = 0.599). In the present study, the HBM construct demonstrated acceptable psychometric properties, with internal consistency reliability (α = 0.717; CR = 0.900) and adequate convergent validity (AVE = 0.599).

### Data analysis

Data were analyzed using SmartPLS (version 4.1) following established guidelines for Partial Least Squares Structural Equation Modeling (PLS-SEM) [60]. This approach is suitable for modeling complex relationships among latent constructs and for estimating direct, indirect (mediation), and moderation effects. Prior to analysis, the data were screened for missing values and multivariate outliers using Mahalanobis distance. Descriptive statistics (means, standard deviations, and ranges) were computed to summarize the study variables.

The analysis was conducted in two stages following established guidelines for partial least squares structural equation modeling (PLS-SEM) (Hair et al., 2017; Hair et al., 2019)[60–61]. First, the measurement model was evaluated in terms of indicator reliability (outer loadings), internal consistency reliability (Cronbach’s alpha, rho_A, and composite reliability), convergent validity (average variance extracted), and discriminant validity using the Fornell–Larcker criterion [62], Heterotrait–Monotrait ratio (HTMT) Henseler et al [63], and cross-loadings. Second, the structural model was assessed using bootstrapping with 10,000 resamples to estimate path coefficients and their statistical significance. Mediation (experiential avoidance) and moderation (age range and sex) effects were examined within the structural model, while multicollinearity was evaluated using variance inflation factor (VIF).

Model predictive performance was assessed using Stone–Geisser’s Q², PLSpredict, and the cross-validated predictive ability test (CVPAT), providing evidence of out-of-sample predictive relevance. Additionally, k-means cluster analysis was conducted using SPSS to identify distinct subgroups based on PTSD symptom profiles, complementing the variable-centered PLS-SEM approach with a person-centered perspective.

## Result

### Assumption Testing and Data Screening

Prior to structural model analysis, the dataset was screened to ensure suitability for partial least squares structural equation modeling (PLS-SEM). Missing data were minimal and appropriately handled using standard statistical procedures. Multivariate outliers were assessed using Mahalanobis distance, and no significant outliers were identified in the dataset. Common method bias was examined, and no substantial bias was detected in the data.

Descriptive statistics for the study variables are presented in Table 1. Perceived susceptibility (M = 33.12, SD = 6.06), perceived severity (M = 23.90, SD = 5.22), and perceived barriers (M = 44.91, SD = 7.33) were observed at moderate levels. Experiential avoidance showed a mean score of 14.43 (SD = 8.04), indicating variability in experiential avoidance across participants. PTSD symptoms had a mean score of 30.34 (SD = 12.05), with subscale means of 9.39 (SD = 4.36) for re-experiencing, 11.16 (SD = 4.90) for avoidance, and 9.79 (SD = 4.21) for hyperarousal.

**Table 1:**
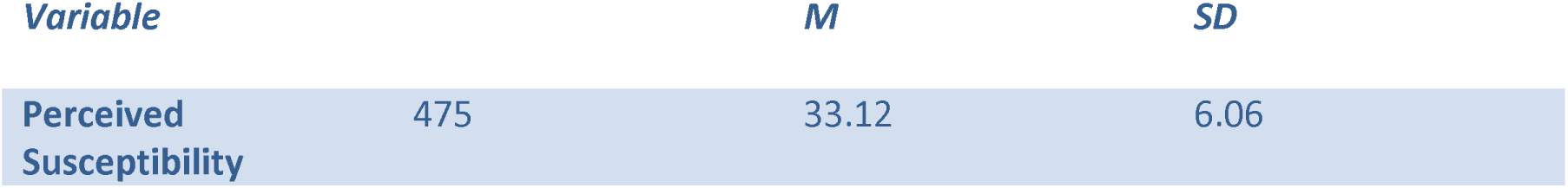
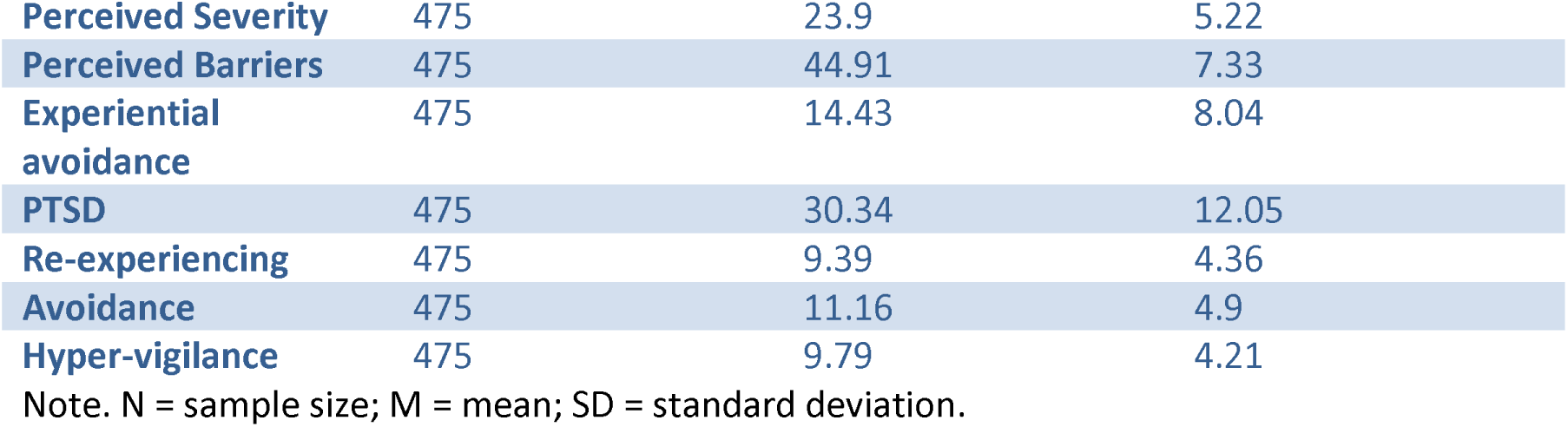
Descriptive Statistics for Key Variables.

| *Variable* |  | *M* | *SD* |
| --- | --- | --- | --- |
| Perceived Susceptibility | 475 | 33.12 | 6.06 |
| **Perceived Severity** | 475 | 23.9 | 5.22 |
| **Perceived Barriers** | 475 | 44.91 | 7.33 |
| **Experiential avoidance** | 475 | 14.43 | 8.04 |
| **PTSD** | 475 | 30.34 | 12.05 |
| **Re-experiencing** | 475 | 9.39 | 4.36 |
| **Avoidance** | 475 | 11.16 | 4.9 |
| **Hyper-vigilance** | 475 | 9.79 | 4.21 |

Note. N = sample size; M = mean; SD = standard deviation.

In terms of distribution, perceived susceptibility was categorized as low (2.9%), moderate (49.5%), and high (47.6%), while perceived severity was predominantly moderate (68.8%). Perceived barriers were distributed as low (15.2%), moderate (50.1%), and high (34.7%). Experiential avoidance scores ranged from 7 to 49, reflecting variability across participants.

### PTSD Prevalence, Symptom Profile, and Clustering

Based on DSM-IV symptom criteria applied to the PTSD Symptom Scale–Civilian Version, the prevalence of PTSD in the sample was 21.9%. Symptom profile analysis indicated that hyperarousal symptoms were most frequently endorsed (81.7%), followed by avoidance (74.2%) and re-experiencing symptoms (68.4%), suggesting that arousal-related symptoms were the most prominent feature of PTSD in the study population.

K-means cluster analysis (k = 2) was conducted to identify heterogeneity in symptom presentation. The analysis revealed two distinct groups comprising a low symptom cluster (66.3%) and a high symptom cluster (33.7%), with clear separation across PTSD indicators based on cluster centroids. One-way ANOVA results confirmed statistically significant differences across all PTSD symptom dimensions (p < .001), indicating meaningful variation in symptom severity between clusters.

### 4.5 Cluster Validation and Group Differences

Independent samples t-tests were conducted to examine differences in Health Belief Model constructs, experiential avoidance, and demographic variables between the two clusters (Table 2). Results indicated that perceived barriers and experiential avoidance significantly differed between groups, while perceived susceptibility and perceived severity did not show statistically significant differences. Among demographic variables, sex showed a significant difference, while age was marginal. Overall, cluster membership was primarily differentiated by experiential avoidance and perceived barriers rather than general risk perception.

**Table 2:** Differences between PTSD Symptom Clusters on Study Variables.

| Variable | Low PTSD<br>(Cluster 1) M ± | High PTSD<br>(Cluster 2) M ± | t | p | Mean<br>Difference | 95% CI |
| --- | --- | --- | --- | --- | --- | --- |

|  | SD (n = 160) | SD (n = 315) |  |  |  |  |
| --- | --- | --- | --- | --- | --- | --- |
| Perceived susceptibility | 32.88 ± 6.19 | 33.25 ± 5.99 | -0.639 | .523 | -0.376 | [-1.532, 0.781] |
| Perceived severity | 23.34 ± 5.34 | 24.19 ± 5.15 | -1.679 | .094 | -0.850 | [-1.844, 0.144] |
| Perceived barriers | 43.12 ± 7.04 | 45.82 ± 7.32 | -3.852 | < .001 | -2.703 | [-4.083, -1.324] |
| Experiential avoidance | 19.66 ± 8.53 | 11.77 ± 6.30 | 11.403 | < .001 | 7.891 | [6.531, 9.251] |
| Age range | 2.78 ± 1.61 | 3.11 ± 1.79 | -1.940 | .053 | -0.327 | [-0.658, 0.004] |
| Sex | 1.79 ± 0.41 | 1.57 ± 0.50 | 4.882 | < .001 | 0.222 | [0.133, 0.312] |

**Note.** Cluster 1 = low PTSD symptom group; Cluster 2 = high PTSD symptom group. Values are presented as mean ± standard deviation. t = independent samples t-test statistic; CI = confidence interval. \*p < .05, \*\*p < .01, \*\*\*p < .001.

As shown in Table 2, to examine differences between the identified PTSD symptom clusters, independent samples t-tests were conducted on Health Belief Model constructs, experiential avoidance, and demographic variables. The results (Table 2) indicate that cluster membership was primarily differentiated by perceived barriers and experiential avoidance, both of which were significantly higher in the high symptom cluster. In contrast, perceived susceptibility and perceived severity did not significantly differ between clusters. Among demographic variables, sex showed a significant difference, while age was marginal.

### Measurement Model Assessment

As shown in Table 3, the measurement model was evaluated using SmartPLS in accordance with established guidelines for partial least squares structural equation modeling [60–61]. The assessment included indicator reliability, internal consistency reliability, convergent validity, and discriminant validity.

**Table 3:** Measurement Model Results (Factor Loadings, Reliability, and Convergent Validity)

| Construct | Indicator | Loading | $\alpha$ | rho_a | CR | AVE |
| --- | --- | --- | --- | --- | --- | --- |
| **Experiential Avoidance** | D38 | 0.658 | 0.852 | 0.857 | 0.888 | 0.532 |
|  | D39 | 0.739 |  |  |  |  |
|  | D40 | 0.778 |  |  |  |  |
|  | D41 | 0.698 |  |  |  |  |
|  | D42 | 0.794 |  |  |  |  |
|  | D43 | 0.738 |  |  |  |  |
|  | D44 | 0.690 |  |  |  |  |
| **Health Belief Model** | Barriers | 0.923 | 0.717 | 0.900 | 0.812 | 0.599 |
|  | Severity | 0.791 |  |  |  |  |
|  | Susceptibility | 0.564 |  |  |  |  |
| **PTSD** | Avoidance | 0.907 | 0.875 | 0.879 | 0.923 | 0.800 |
|  | Hyper-vigilance | 0.884 |  |  |  |  |
|  | Re-experiencing | 0.892 |  |  |  |  |

Note: $\alpha$ = Cronbach's alpha; $\rho_A$ = Dijkstra–Henseler's rho; CR = composite reliability; AVE = average variance extracted.

Indicator reliability was assessed using outer loadings. Most indicators exceeded the recommended threshold of 0.70, indicating adequate item reliability. However, a few indicators showed slightly lower loadings, including perceived susceptibility (0.564) within the Health Belief Model and selected experiential avoidance items (D38 = 0.658; D41 = 0.698; D44 = 0.690). These indicators were retained based on theoretical relevance, as their removal did not result in substantial improvements in composite reliability or average variance extracted [60].

Internal consistency reliability was assessed using Cronbach’s alpha, rho_A, and composite reliability (CR). All constructs demonstrated acceptable to strong reliability. Although Cronbach’s alpha remains widely used in behavioral research, composite reliability and rho_A provide more robust estimates of internal consistency, as Cronbach’s alpha assumes tau-equivalence among indicators, which is often unrealistic in applied research contexts.

Convergent validity was assessed using Average Variance Extracted (AVE). All constructs exceeded the recommended threshold of 0.50, indicating adequate convergent validity. Experiential avoidance (AVE = 0.52), the Health Belief Model (AVE = 0.66), and PTSD (AVE = 0.77) all demonstrated satisfactory levels of explained variance.

As demonstrated in table 4, discriminant validity was assessed using the Heterotrait–Monotrait ratio (HTMT) and the Fornell–Larcker criterion. As shown in Table [2], all HTMT values were below the conservative threshold of 0.85, confirming adequate discriminant validity. In addition, the Fornell–Larcker criterion was satisfied, as the square roots of AVE exceeded inter-construct correlations for all constructs [62].

**Table 4:** Discriminant Validity Heterotriat moontriat ratio.

| Construct | EA | HBM | PTSD |
| --- | --- | --- | --- |
| **EA** | — | 0.188 | 0.508 |
| **HBM** | — | — | 0.196 |
| **PTSD** | — | — | — |

**Note:** HTMT values below 0.85 indicate adequate discriminant validity.

### Structural model assessment

The structural model in Figure 1 was assessed using Partial Least Squares Structural Equation Modeling (PLS-SEM) in SmartPLS to estimate the relationships among latent constructs and test the proposed hypotheses. Bootstrapping with 10,000 resamples was conducted to evaluate the significance of path coefficients and to generate bias-corrected confidence intervals. This non-parametric procedure was used to assess the stability and robustness of the estimated effects.

**Figure 1:**
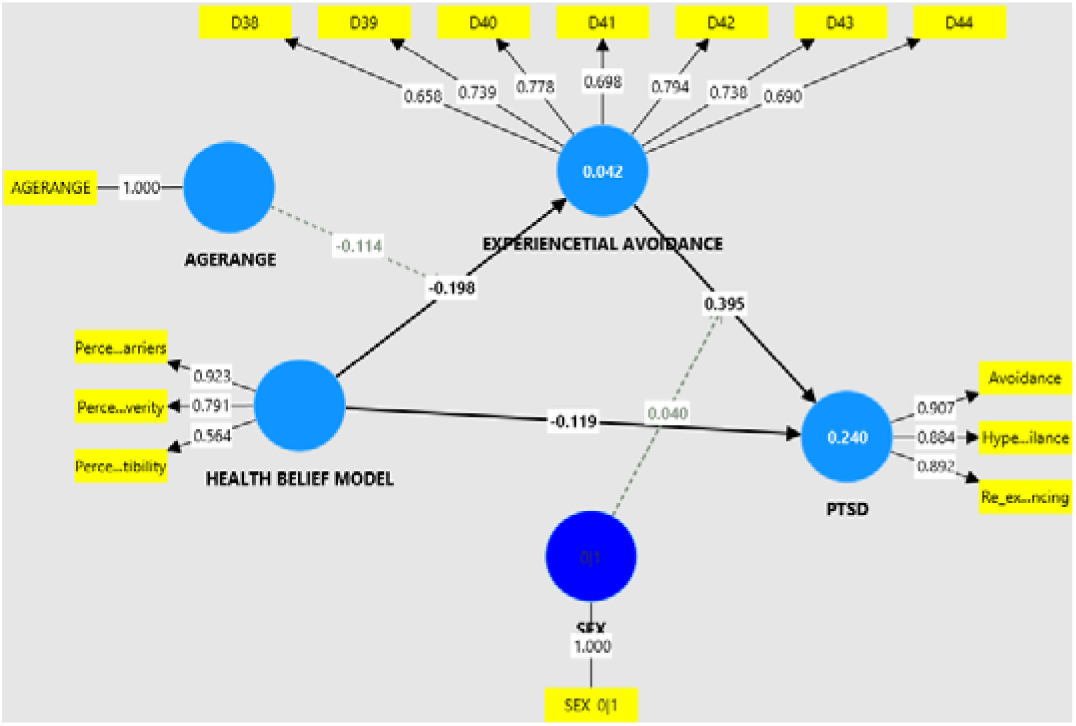
Structural Model Showing Path Coefficients and Relationships. Note. Values represent standardized path coefficients (β) obtained from bootstrapping (10,000 resamples). *p* < .05, ***p*** < .01, ***p*** < .001. EA = Experiential Avoidance; HBM = Health Belief Model; PTSD = Post-Traumatic Stress Disorder.

Mediation effects were examined by assessing the significance of specific indirect effects using bootstrapped confidence intervals, with mediation established when the confidence intervals did not include zero. Moderation effects were tested through the inclusion of interaction terms in the structural model, and their significance was evaluated using bootstrapping. jMulticollinearity was assessed using variance inflation factor (VIF). All VIF values ranged from 1.000 to 2.440, which are well below the recommended threshold of 5.0 (Hair et al [61] and the more conservative threshold of 3.3 Kock [64], indicating that multicollinearity was not a concern among the predictor constructs and interaction terms in the model (see figure 1).

### Model Fit

The model fit was assessed using the Standardized Root Mean Square Residual (SRMR), d_ULS, d_G, Chi-square, and Normed Fit Index (NFI). The SRMR values for both the saturated model (0.063) and the estimated model (0.073) were below the recommended threshold of 0.08, indicating an acceptable model fit.

Although the NFI values (0.786 for the saturated model and 0.640 for the estimated model) were below the conventional cut-off of 0.90, this is common in PLS-SEM, where model fit indices are considered secondary to predictive accuracy and explanatory power. The d_ULS and d_G values (0.484–0.641 and 0.188–0.294, respectively) further support acceptable model fit, although these indices are primarily used for model comparison rather than absolute evaluation.

Overall, the results indicate that the structural model demonstrates an acceptable level of model fit based on SRMR, supporting its suitability for hypothesis testing.

### 4.5 Predictive Relevance and Predictive Performance

The predictive capability of the structural model was assessed using both in-sample and out-of-sample procedures. Predictive relevance was first evaluated using blindfolding based on cross-validated redundancy (Q²). The results indicate that PTSD demonstrated moderate predictive relevance (Q² = 0.144), suggesting that the model captures a meaningful proportion of variance in the primary endogenous construct. In contrast, experiential avoidance showed a small Q² value (Q² = 0.016), indicating weak predictive relevance. As expected, exogenous variables such as age, sex, and Health Belief Model (HBM) constructs yielded Q² values of zero, as they are not predicted within the structural model.

At the indicator level, PTSD-related dimensions exhibited relatively stronger predictive relevance, particularly avoidance (Q² = 0.171), re-experiencing (Q² = 0.134), and hypervigilance (Q² = 0.127), whereas several other indicators showed low Q² values, suggesting limited predictive contribution at the item level.

To further assess out-of-sample predictive performance, PLSpredict and cross-validated predictive ability testing (CVPAT) were conducted as supplementary diagnostic procedures. The PLSpredict results indicated limited predictive performance, as Q²predict values were close to zero and root mean squared error (RMSE) values showed minimal improvement over linear model benchmarks. Similarly, CVPAT results indicated that the PLS-SEM model did not consistently outperform the linear benchmark model across constructs.

Taken together, these findings suggest that the model demonstrates stronger explanatory relevance than predictive performance, with moderate in-sample predictive relevance for PTSD but limited out-of-sample predictive accuracy. This pattern is consistent with the primary aim of the study, which is to explain the cognitive–emotional mechanisms linking Health Belief Model constructs, experiential avoidance, and PTSD symptoms rather than to optimize prediction.

### Structural Model Results

As indicated in Table 5, the structural model was assessed using bootstrapping with 10,000 resamples to evaluate the hypothesized direct, indirect, moderating, and conditional indirect relationships among Health Belief Model (HBM) constructs, experiential avoidance, demographic variables, and PTSD symptoms.

**Table 5:**
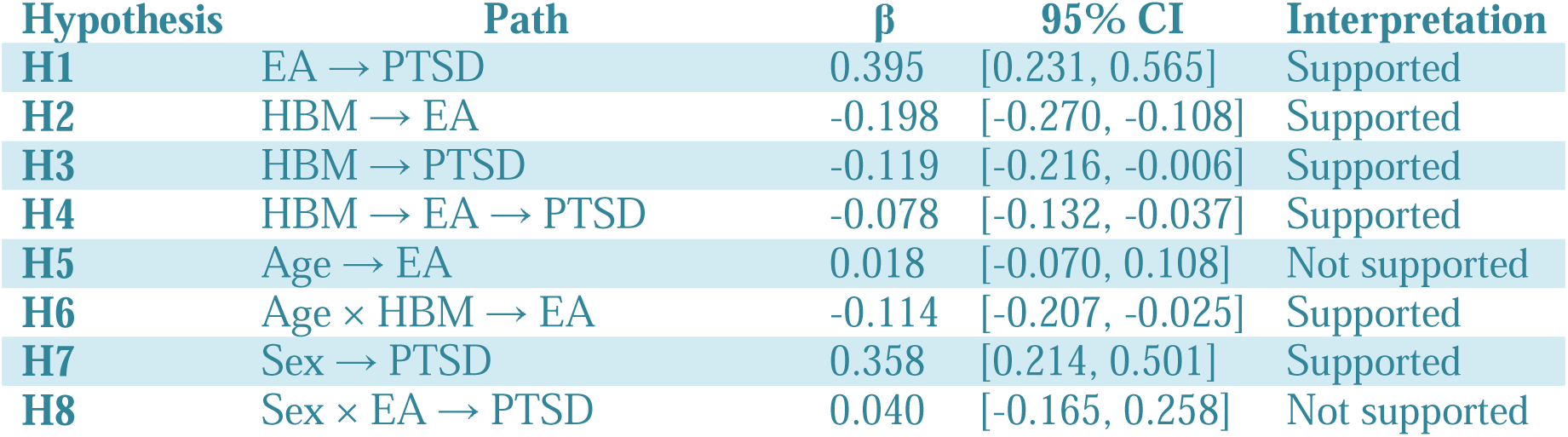
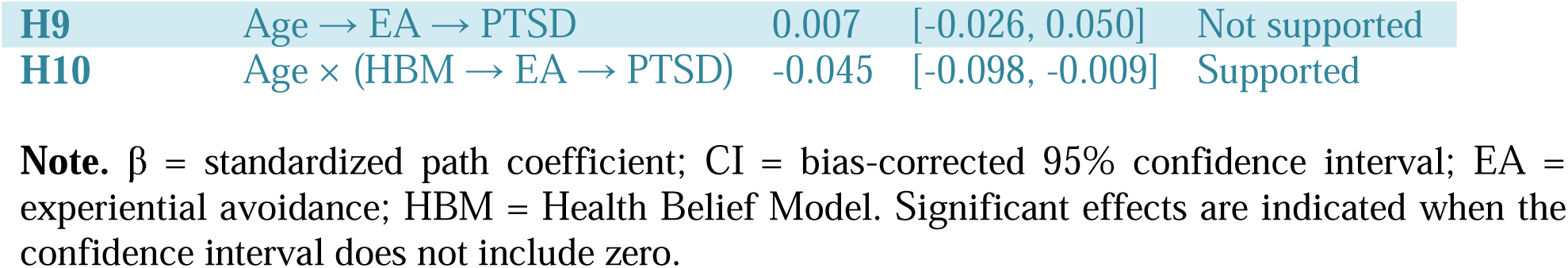
Structural Model Results (Bootstrapped Path Coefficients)

Experiential avoidance was positively associated with PTSD symptoms (β = 0.395, 95% CI [0.231, 0.565]), supporting H1. Health Belief Model (HBM) constructs were negatively associated with experiential avoidance (β = −0.198, 95% CI [−0.270, −0.108]), supporting H2, and negatively associated with PTSD symptoms (β = −0.119, 95% CI [−0.216, −0.006]), supporting H3.

The indirect effect of HBM on PTSD symptoms through experiential avoidance was significant (β = −0.078, 95% CI [−0.132, −0.037]), supporting H4 and indicating partial mediation. By combining the direct and indirect pathways, the total effect of HBM on PTSD symptoms was estimated at β = −0.197, indicating that stronger health beliefs were associated with lower PTSD symptoms through both direct and indirect pathways.

Age was not significantly associated with experiential avoidance (β = 0.018, 95% CI [−0.070, 0.108]), and therefore H5 was not supported. However, age significantly moderated the relationship between HBM constructs and experiential avoidance (β = −0.114, 95% CI [−0.207, −0.025]), supporting H6. As seems in a simple slop analysis in figure 2, at low levels of health beliefs (−1 SD), experiential avoidance is highest among older individuals (≈ +0.30) compared to younger individuals (≈ +0.05). However, at high levels of health beliefs (+1 SD), experiential avoidance decreases substantially among older individuals (≈ −0.30) but only slightly among younger individuals (≈ −0.10). This indicates that the negative association between health beliefs and experiential avoidance is markedly stronger in older individuals.

**Figure 2.**
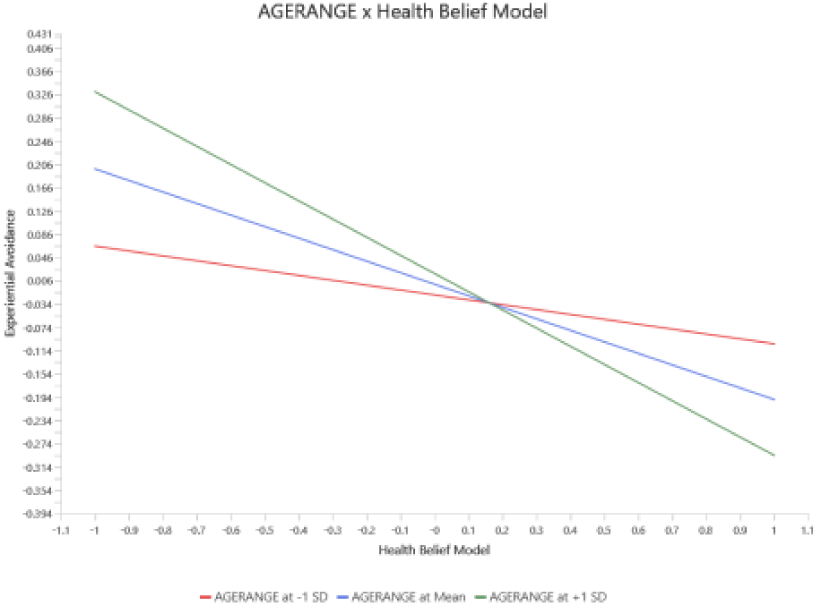
goes here: Age × HBM → Experiential Avoidance interaction plot. **Note.** The figure illustrates the interaction between Age Range and Health Belief Model (HBM) on Experiential Avoidance at low (−1 SD), mean, and high (+1 SD) levels of Age Range.

**Figure 3.**
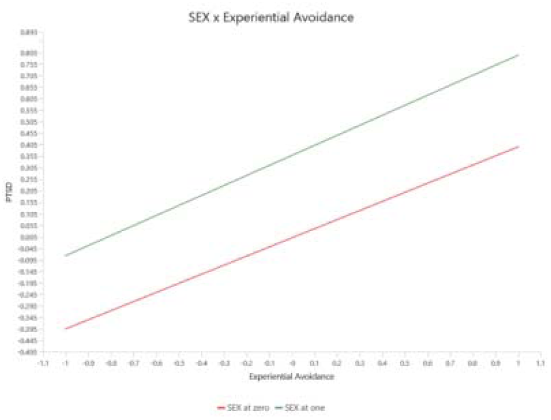
goes here: Sex × Experiential Avoidance → PTSD interaction plot. Note. The figure illustrates the interaction between Sex and Experiential Avoidance in predicting PTSD symptoms at low (−1 SD) and high (+1 SD) levels of Experiential Avoidance..

However, the moderating effect of sex on the relationship between experiential avoidance and Sex showed a significant positive association with PTSD symptoms (β = 0.358, 95% CI [0.214, 0.501]), supporting H7. However, the moderating effect of sex on the relationship between experiential avoidance and PTSD symptoms was not significant (β = 0.040, 95% CI [−0.165, 0.258]), and therefore H8 was not supported. Experiential avoidance (EA) showed a consistent positive relationship with PTSD symptoms across both groups, increasing from approximately **−0.35 to 0.45** in the lower group and from **0.00 to 0.80** in the higher group as EA moved from −1 SD to +1 SD. The higher group consistently exhibited greater PTSD symptoms across all levels of EA, with a stable between-group difference and parallel slopes, indicating no meaningful Sex × EA interaction effect.

The indirect effect of age on PTSD symptoms through experiential avoidance was not significant (β = 0.007, 95% CI [−0.026, 0.050]), indicating that H9 was not supported. In contrast, the conditional indirect effect of age on the relationship between HBM constructs and PTSD symptoms through experiential avoidance was significant (β = −0.045, 95% CI [−0.098, −0.009]), supporting H10 and indicating moderated mediation.

Overall, the findings suggest that experiential avoidance functions as an important psychological pathway linking health beliefs to PTSD symptoms. Health beliefs were associated with lower experiential avoidance and reduced PTSD symptoms, while age conditioned the strength of the indirect relationship between HBM constructs and PTSD symptoms through experiential avoidance.

## DISCUSSION

This study examined a cognitive–emotional framework in which health beliefs influence post-traumatic stress disorder (PTSD) symptoms both directly and indirectly through experiential avoidance, while demographic factors condition these relationships. Overall, the findings provide partial support for this model, highlighting the joint role of cognitive appraisals and emotion regulation processes in shaping PTSD symptom severity among healthcare workers.

Experiential avoidance emerged as a key mechanism linking health beliefs to PTSD, while demographic factors played differentiated roles, with age acting as a contextual moderator and sex directly associated with PTSD symptoms. Together, these findings reflect the direct, mediating, and conditional pathways specified in the study.

Experiential avoidance emerged as the strongest predictor of PTSD symptoms (β = 0.395, 95% CI [0.231, 0.565]), supporting process-based models of psychopathology and theoretical frameworks such as Acceptance and Commitment Therapy, which conceptualize avoidance of internal experiences as a central mechanism underlying psychological distress [9,28]. Healthcare workers reporting higher levels of experiential avoidance also demonstrated greater PTSD symptom severity, suggesting that avoidance-based coping may be maladaptive in high-stress clinical environments during infectious disease outbreaks. In healthcare settings such as those in Ekiti State, repeated exposure to emotionally demanding situations may increase reliance on avoidance-oriented coping strategies.

This interpretation was further supported by the cluster analysis, which demonstrated substantially higher levels of experiential avoidance among participants in the high PTSD symptom cluster compared with the low symptom cluster. The convergence between the structural model and person-centered findings suggests that experiential avoidance may represent an important marker of PTSD symptom severity among healthcare workers [17].

Health Belief Model constructs were negatively associated with both experiential avoidance (β = −0.198, 95% CI [−0.270, −0.108]) and PTSD symptoms (β = −0.119, 95% CI [−0.216, −0.006]), indicating that more adaptive health-related cognitive appraisals were linked to lower psychological distress. This finding is consistent with the Health Belief Model, which proposes that perceptions of susceptibility, severity, and barriers shape emotional and behavioral responses to health threats [26,27]. Additional insight from the cluster analysis revealed that perceived barriers differed significantly across PTSD symptom groups, with higher levels observed in the high PTSD cluster, whereas perceived susceptibility and severity did not significantly differ between groups. This finding is consistent with previous studies suggesting that perceived barriers may be more closely associated with psychological distress than general perceptions of health risk [65].

The indirect effect of Health Belief Model constructs on PTSD symptoms through experiential avoidance was significant (β = −0.078, 95% CI [−0.132, −0.037]), suggesting that avoidance-based coping may represent an important psychological mechanism linking health-related cognitions to trauma-related distress. Healthcare workers with more adaptive perceptions of health threats may have been less likely to rely on experiential avoidance when confronted with emotionally distressing situations, thereby reducing vulnerability to PTSD symptoms. This interpretation is consistent with cognitive–behavioral and process-based frameworks, which propose that maladaptive avoidance of internal experiences may maintain or intensify psychological distress over time [15,30–32].

In the context of the COVID-19 pandemic, healthcare workers were repeatedly exposed to uncertainty, fear of infection, high workloads, and emotionally demanding clinical conditions. Under such circumstances, adaptive health beliefs may facilitate more effective cognitive and emotional processing of stressors, thereby reducing disengagement from distressing thoughts and emotions. Collectively, the findings suggest that experiential avoidance may partially explain how health-related beliefs translate into psychological outcomes among frontline healthcare workers.

Taken together, the direct and indirect pathways indicate that Health Belief Model constructs exerted an overall negative effect on PTSD symptoms (total effect β = −0.197), suggesting that adaptive health beliefs were broadly associated with lower psychological distress among healthcare workers. Although previous studies have examined Health Belief Model constructs in relation to health behaviors and emotional responses to health threats, the present findings extend this literature by demonstrating both direct and indirect associations between health beliefs and PTSD symptoms through experiential avoidance.

The mediation analysis further indicated that experiential avoidance partially accounted for this association, consistent with a partial mediation pattern. Although the cluster analysis did not directly examine mediation effects, the finding that both experiential avoidance and perceived barriers differentiated PTSD symptom groups provides additional support for the proposed cognitive–emotional framework.

Demographic variables appeared to operate as boundary conditions rather than simple predictors. Although age was not significantly associated with experiential avoidance, age significantly moderated both the association between Health Belief Model constructs and experiential avoidance and the indirect pathway linking Health Belief Model constructs to PTSD symptoms through experiential avoidance. These findings suggest that age may influence the strength of the cognitive–emotional processes underlying psychological distress rather than directly influencing avoidance responses themselves.

In contrast, sex showed a direct association with PTSD symptoms, with higher levels observed in one group. This finding aligns with prior research showing higher PTSD symptom severity among female healthcare workers [48–50]. However, sex did not moderate the relationship between experiential avoidance and PTSD, indicating that the association between experiential avoidance and PTSD is consistent across genders.

In summary, these findings extend the Health Belief Model by demonstrating its relevance not only to health behavior but also to trauma-related psychological outcomes. When considered alongside emotion regulation frameworks, the results highlight experiential avoidance as a key process linking cognitive appraisals to PTSD symptoms. Furthermore, the findings indicate that demographic factors such as age and sex do not fundamentally alter these relationships but instead are associated with variation in their strength. This integrative perspective provides a more comprehensive understanding of how cognitive and emotional processes are related to trauma-related distress among healthcare workers in high-risk environments.

### Conclusion

This study demonstrates that cognitive health beliefs and experiential avoidance operate within a conditional cognitive–emotional framework to contribute to PTSD symptoms among healthcare workers. The findings highlight experiential avoidance as a key psychological process linking health-related appraisals to trauma-related distress, while perceived barriers emerged as the most influential cognitive component of the Health Belief Model. Demographic factors shaped the strength of these relationships rather than their structural form, with sex showing a direct association with PTSD symptoms and age moderating the cognitive–emotional pathway. Specifically, age influenced the strength of the relationship between health beliefs and experiential avoidance, as well as the indirect pathway to PTSD.

Although the model demonstrated limited out-of-sample predictive power relative to a linear benchmark, it provides meaningful explanatory insight into the cognitive and emotional mechanisms underlying PTSD in this population.Overall, the findings underscore the importance of integrating cognitive appraisal and emotion regulation frameworks in understanding trauma-related distress and suggest that interventions targeting experiential avoidance and perceived barriers may be beneficial in high-risk healthcare settings.

## THEORETICAL IMPLICATIONS

This study extends the Health Belief Model (HBM) beyond its traditional focus on health-related behaviors by demonstrating its relevance to trauma-related psychological outcomes among healthcare workers. The findings indicate that cognitive appraisals of health threats are associated with PTSD symptoms through both direct and indirect pathways, suggesting that the model has explanatory value beyond preventive health decision-making.

An integrated cognitive–emotional interpretation of the results further suggests that experiential avoidance functions as a key mechanism through which health beliefs translate into psychological distress. This implies that PTSD is not solely a function of cognitive evaluation of risk, but is also shaped by how individuals regulate the emotional responses generated by such appraisals. In this sense, cognitive and emotional processes operate interactively rather than independently in explaining trauma-related outcomes. The findings also refine the Health Belief Model by indicating that its components do not exert uniform influence in psychological contexts. Among the constructs examined, perceived barriers emerged as the most consistent predictor of distress, suggesting that structural and contextual constraints may play a more proximal role in shaping PTSD symptoms than more abstract risk perceptions such as susceptibility and severity. This pattern implies that future theoretical applications of the HBM in mental health contexts may benefit from considering differential weighting of its components.

Further theoretical insight is provided by the cluster analysis, which revealed distinct patterns of PTSD symptom expression. The clear separation between high and low symptom groups, particularly along dimensions of experiential avoidance and perceived barriers, suggests that PTSD in this population is better understood as a heterogeneous and process-based condition rather than a uniform response to trauma exposure. Finally, demographic variables appear to function as contextual boundary conditions within this framework. Age influences the strength of the cognitive–emotional pathway linking health beliefs and experiential avoidance, while sex is directly associated with PTSD symptoms without altering the underlying psychological mechanism. This suggests that demographic characteristics shape the intensity of psychological processes rather than defining their structure.

## LIMITATIONS

Despite its contributions, this study has several limitations that should be considered when interpreting the findings. The cross-sectional design limits the ability to draw causal inferences, as the temporal ordering of relationships among cognitive beliefs, experiential avoidance, and PTSD symptoms cannot be established. Longitudinal research is therefore needed to clarify the direction and stability of these associations over time.

In addition, the use of self-reported measures introduces the possibility of response biases, including social desirability and recall bias. This is particularly relevant in studies involving psychological distress among healthcare workers, where reporting may be influenced by occupational concerns. Future studies could strengthen validity by incorporating clinical interviews or behavioral indicators alongside self-report measures. Although the measurement model demonstrated acceptable reliability and validity, some Health Belief Model indicators, particularly perceived susceptibility, showed relatively weaker loadings. This suggests that the operationalization of certain constructs may require refinement in future research to ensure stronger construct representation in similar contexts.

The study was also limited to healthcare workers in Ekiti State, which may restrict the generalizability of the findings to other regions or healthcare systems with different organizational structures, resources, and stress exposures. Replication in diverse settings is therefore necessary. While cluster analysis provided useful evidence of heterogeneity in PTSD symptom severity, it is a data-driven approach and does not imply causal classification. As such, it should be interpreted as complementary to the structural model rather than evidence of latent causal groupings. Finally, although PLS-SEM is well suited for complex predictive–explanatory models, its predictive assessment procedures indicated limited out-of-sample performance relative to simpler benchmarks. In addition, PLS-SEM prioritizes variance explanation rather than global model fit, which limits direct comparison with covariance-based structural equation modeling approaches.

## FUTURE RESEARCH DIRECTIONS

Future research should adopt longitudinal designs to better examine the temporal dynamics among cognitive health beliefs, experiential avoidance, and PTSD symptoms. Such designs would help clarify the directionality of effects and provide stronger evidence regarding how these psychological processes evolve under sustained occupational stress. Further investigation is also needed to unpack the differential roles of Health Belief Model components. The present findings suggest that perceived barriers may be more strongly associated with psychological distress than perceived susceptibility and severity, raising questions about whether certain components exert more proximal influence in mental health applications of the model.

In addition, intervention-based studies are needed to determine whether reductions in experiential avoidance translate into meaningful improvements in PTSD symptoms among healthcare workers. This would help establish the clinical relevance of targeting emotion regulation processes within trauma-informed interventions. Future research should also incorporate broader contextual and individual difference variables, such as resilience, workload, organizational support, coping styles, and prior trauma exposure. These factors may further clarify vulnerability and protective mechanisms in high-stress healthcare environments. Given the heterogeneous PTSD profiles observed in this study, future work should employ advanced person-centered approaches such as latent profile analysis or mixture modeling to further validate and extend the cluster-based findings and improve understanding of psychological subgroups.

Moreover, future studies should evaluate the predictive performance of the proposed model using longitudinal or external datasets to determine its generalizability beyond cross-sectional estimation and strengthen its applied relevance.

Finally, qualitative approaches would provide deeper insight into how healthcare workers interpret health threats and regulate emotional responses in real-world clinical settings, thereby complementing quantitative findings and enhancing theoretical development.

## Data Availability

All data produced in the present study are available upon reasonable request to the authors

https://github.com/jamboyar/covid-19/blob/main/HEALTH%20BELIEF%20DATA.sav

## Data availability

The data used in this study are available from the corresponding author upon reasonable request.

## References

1. Huang, Y., & Zhao, N. (2020). Generalized anxiety disorder, depressive symptoms and sleep quality during COVID-19 outbreak in China: a web-based cross-sectional survey. Psychiatry research, 288, 112954. 10.1016/j.psychres.2020.112954

2. Rathore FA, Farooq F. Citations from predatory journals must be discouraged and how to identify predatory journals and publishers. Ir J Med Sci [Internet]. 2021 Nov;190(4):1645–1646 [cited 2026 Apr 26]. Available from: https://pmc.ncbi.nlm.nih.gov/articles/PMC7410455

3. African Union and Africa CDC, 2020. Coronavirus Disease 2019 (COVID-19). Latest Updates on the COVID-19 Crisis from Africa CDC. Available at: https://africacdc.org/covid-19/. Accessed May 26, 2020. [<u>Google Scholar</u>]

4. World Health Organization. Country health system profile: Nigeria [Internet]. Geneva: World Health Organization; 2024 [cited 2026 Apr 26]. Available from:

5. deloye D, David RA, Olaogun AA, et al. Health workforce and governance: the crisis in Nigeria. Hum Resour Health 2017;15:32. 10.1186/s12960-017-0205-4 [DOI] [PMC free article] [PubMed] [Google Scholar]

6. 4.World Health Organisation. Global health workforce statistics database, 2022. Available: https://www.who.int/data/gho/data/themes/topics/health-workforce [Accessed 30 June 2022].

7. Carmassi C, Foghi C, Dell’Oste V, Cordone A, Bertelloni CA, Bui E, et al. PTSD symptoms in healthcare workers facing the three coronavirus outbreaks: What can we expect after the COVID-19 pandemic. Psychiatry Res. (2020) 292:113312. doi: 10.1016/j.psychres.2020.113312

8. Blekas A, Voitsidis P, Athanasiadou M, Parlapani E, Chatzigeorgiou AF, Skoupra M, et al. COVID-19: PTSD symptoms in Greek health care professionals. Psychol Trauma Theory Res Pract Policy. (2020) 12:812– 9. doi: 10.1037/tra0000914

9. Hayes-Skelton SA, Eustis EH. Experiential avoidance. In: Abramowitz JS, Blakey SM, editors. Clinical handbook of fear and anxiety: Maintenance processes and treatment mechanisms. American Psychological Association; 2020. p. 115–131. doi: 10.1037/0000150-007.

10. Chen H, Li X, Gao J, Liu X, Mao Y, Wang R, Zheng P, Xiao Q, Jia Y, Fu H, Dai J Health Belief Model Perspective on the Control of COVID-19 Vaccine Hesitancy and the Promotion of Vaccination in China: Web-Based Cross-sectional Study J Med Internet Res 2021;23(9):e29329 doi: 10.2196/29329

11. Wong LP, Alias H, Wong PF, Lee HY, AbuBakar S. The use of the health belief model to assess predictors of intent to receive the COVID-19 vaccine and willingness to pay. Hum Vaccin Immunother. 2020;16(9):2204–2214. doi:10.1080/21645515.2020.1790279.

12. Zhang Y, Li X, Wang L, et al. Psychological resilience and mental health outcomes among healthcare workers during the COVID-19 pandemic: a cross-sectional study. Front Psychiatry [Internet]. 2024;15:1323111 [cited 2026 Apr 27]. Available from: 10.3389/fpsyt.2024.1323111

13. Sahebi A, Yousefi A, Abdi K, Jamshidbeigi Y, Moayedi S, Torres M, et al. The prevalence of post-traumatic stress disorder among health care workers during the COVID-19 pandemic: an umbrella review and meta-analysis. Front Psychiatry. 2021;12:764738. doi:10.3389/fpsyt.2021.764738.

14. North, C.S.; Surís, A.M.; Pollio, D.E. A Nosological Exploration of PTSD and Trauma in Disaster Mental Health and Implications for the COVID-19 Pandemic. Behav. Sci. 2021, 11, 7. doi:10.3390/bs11010007

15. Hayes-Skelton SA, Eustis EH. Experiential avoidance. In: Abramowitz JS, Blakey SM, editors. Clinical handbook of fear and anxiety: Maintenance processes and treatment mechanisms. American Psychological Association; 2020. p. 115–131. doi: 10.1037/0000150-007.

16. Wang Y, Tian J, Yang Q. Experiential avoidance process model: A review of the mechanism for the generation and maintenance of avoidance behavior. Psychiatry Clin Psychopharmacol. 2024;34(2):179–190. doi:10.5152/pcp.2024.23777

17. Yarseah DA, Ezeani ESU, Ogunsanmi JO, Agboola SM, Ogunleye TS, Ahmed AN. The pathway to the development and maintenance of PTSD among healthcare workers in the first five months of the COVID-19 pandemic in Ekiti State, Nigeria. Research Square [Preprint]. 2022. Available from: 10.21203/rs.3.rs-1601068/v1

18. Karekla M, Forsyth JP, Kelly MM. Emotional avoidance and panicogenic responding to a biological challenge procedure. Behav Ther. 2004;35(4):725 746. ( 10.1016/S0005-7894(04)80017-0) [DOI] [Google Scholar

19. Fawzy TI, Hecker JE, Clark J. The relationship between cognitive avoidance and attentional bias for snake-related thoughts. J Anxiety Disord. 2006;20(8):1103 1117. ( 10.1016/j.janxdis.2006.01.010) [DOI] [PubMed] [Google Scholar]

20. Lavy EH, van den Hout MA. Cognitive avoidance and attentional bias: Causal relationships. Cognit Ther Res. 1994;18(2):179 191. ( 10.1007/BF02357223) [DOI] [Google Scholar]

21. Zettle RD, Hocker TR, Mick KA, et al. Differential strategies in coping with pain as a function of level of experiential avoidance. Psychol Rec. 2005;55(4):511 524. ( 10.1007/BF03395524) [DOI] [Google Scholar]

22. Hagger MS, Cameron LD, Hamilton K, Hankonen N, Lintunen T, editors. The handbook of behavior change. Cambridge: Cambridge University Press; 2020. doi:10.1017/9781108677318

23. Cormier S, Nurius PS, Osborn CJ. Interviewing and change strategies for helpers: fundamental skills and cognitive behavioral interventions. 6th ed. Belmont (CA): Brooks/Cole, Cengage Learning; 2013.

24. Zettle RD, Hocker TR, Mick KA, et al. Differential strategies in coping with pain as a function of level of experiential avoidance. Psychol Rec. 2005;55(4):511 524. ( 10.1007/BF03395524) [DOI] [Google Scholar]

25. Amiri M, Mikal ZM, Sadeghi E, Khosravi A. The prevalence of posttraumatic stress disorder and its correlation with health beliefs among medical students. J Educ Health Promot. 2025 Jan;14(1):25. doi:10.4103/jehp.jehp_1625_23.

26. Janz NK, Becker MH. The Health Belief Model: a decade later. Health Educ Q. 1984 Spring;11(1):1–47. doi:10.1177/109019818401100101.

27. Rosenstock IM, Strecher VJ, Becker MH. Social learning theory and the health belief model. Health Educ Behav. 1988;15(2):175–83. doi:10.1177/109019818801500203

28. Hayes S, Wilson K, Gifford E, Follette V, & Stroshal K (1996). Experiential avoidance and behavioral disorders: A functional dimensional approach to diagnosis and treatment. Journal of Consulting and Clinical Psychology, 64, 1152–1168. doi:10.1037//0022-006X.64.6.1152

29. Chapman AL, Gratz KL, & Brown MZ (2006). Solving the puzzle of deliberate self-harm: The experiential avoidance model. Behaviour Research and Therapy, 44, 371–394. doi:10.1016/j.brat.2005.03.005

30. Hayes S, Strosahl K, & Wilson K (1999). Acceptance and commitment therapy. An experiential approach to behavior change. New York: Guillford

31. Hayes S, Luoma JB, Bond FW, Masuda A, & Lillis J (2006). Acceptance and Commitment Therapy: Model, Process and Outcomes. Behaviour Research and Therapy, 44, 1–25. doi:10.1016/j.brat.2005.06.006

32. Haggeer MS, Cameron LD, Hamilton K, Handkonen N, Lintunen T. Handbook of behavior change. Cambridge: Cambridge University Press; 2020. doi: 10.1017/9781108677218

33. Getachew T, Lami M, Eyeberu A, Balis B, Debella A, Eshetu B, Degefa M, Mesfin S, Negash A, Bekele H, Turiye G, Tamiru D, Nigussie K, Asfaw H, Dessie Y, Alemu A and Sertsu A (2022) Acceptance of COVID-19 vaccine and associated factors among health care workers at public hospitals in Eastern Ethiopia using the health belief model. Front. Public Health 10:957721. doi: 10.3389/fpubh.2022.957721

34. Chen H, Li X, Gao J, Liu X, Mao Y, Wang R, Zheng P, Xiao Q, Jia Y, Fu H, Dai J Health Belief Model Perspective on the Control of COVID-19 Vaccine Hesitancy and the Promotion of Vaccination in China: Web-Based Cross-sectional Study J Med Internet Res 2021;23(9):e29329

35. Wong LP, Alias H, Wong PF, Lee HY, AbuBakar S. The use of the health belief model to assess predictors of intent to receive the COVID-19 vaccine and willingness to pay. Hum Vaccin Immunother. 2020;16(9):2204–2214. doi:10.1080/21645515.2020.1790279.

36. Coe AB, Gatewood SB, Moczygemba LR, Goode JV, Beckner JO. The use of the health belief model to assess predictors of intent to receive the novel (2009) H1N1 influenza vaccine. Innov Pharm. 2012;3(2):1–11. doi:10.24926/iip.v3i2.257.

37. Wong LP, Alias H, Wong PF, Lee HY, AbuBakar S. The use of the health belief model to assess predictors of intent to receive the COVID-19 vaccine and willingness to pay. Hum Vaccin Immunother. 2020;16(9):2204–2214. doi:10.1080/21645515.2020.1790279.

38. Orji R, Vassileva J, Mandryk R. Towards an effective health interventions design: an extension of the Health Belief Model. Online J Public Health Inform. 2012;4(3):e61050. doi:10.5210/ojphi.v4i3.4321. (verify DOI if required)

39. Nguyen K, Vu B, Chandna S, Schultz JH and Mayer G (2025) Between the lines: investigating health beliefs and emotional expressions in online mental health communities. Front. Psychol. 16:1521623. doi: 10.3389/fpsyg.2025.1521623

40. Zhao YC, Zhao M, Song S. Online health information seeking among patients with chronic conditions: integrating the health belief model and social support theory. J Med Internet Res. 2022;24(11):e42447. doi:10.2196/42447.

41. Bond FW, Hayes SC, Baer RA, Carpenter KM, Guenole N, Orcutt HK, et al. Preliminary psychometric properties of the Acceptance and Action Questionnaire–II: A revised measure of psychological inflexibility and experiential avoidance. Behav Ther. 2011;42(4):676–88. doi:10.1016/j.beth.2011.03.007.

42. Menendez-Aller A, Postigo A, Gonzalez-Nuevo C, Gracia-Fernandez J, Gracia-Cueto E. Validation of the Acceptance and Action Questionnaire II in the general Spanish population. Curr Psychol. 2023;42:12096–12103. doi: 10.1007/s12144-021-02447-

43. Charles ST, Hong J. Strength and vulnerability integration. In: Pachana NA, editor. Encyclopedia of Geropsychology. Singapore: Springer; 2015. p. [chapter pages]. doi:10.1007/978-981-287-082-7_10

44. Benson L, English T, Conroy DE, Pincus AL, Gerstorf D, Ram N. Age differences in emotion regulation strategy use, variability, and flexibility: an experience sampling approach. Dev Psychol. 2019 Sep;55(9):1951–1964. doi:10.1037/dev0000727

45. Carstensen LL. Socioemotional selectivity theory: the role of perceived endings in human motivation. Gerontologist. 2021 Oct 27;61(8):1188–1196. doi:10.1093/geront/gnab116

46. Schaufeli WB, Leiter MP, Maslach C. Burnout: 35 years of research and practice. Career Dev Int. (2009) 14:204–20. 10.1108/13620430910966406 [DOI] [Google Scholar]

47. Chou LP, Li CY, Hu SC. Job stress and burnout in hospital employees: comparisons of different medical professions in a regional hospital in Taiwan. BMJ Open. (2014) 4:e004185. 10.1136/bmjopen-2013-004185 [DOI] [PMC free article] [PubMed] [Google Scholar

48. Weinberg A, Creed F. Stress and psychiatric disorder in healthcare professionals and hospital staff. Lancet. (2000) 355:533–7. doi: 10.1016/S0140-6736(99)07366-3

49. Wall TD, Bolden RI, Borrill CS, Carter AJ, Golya DA, Hardy GE, et al. Minor psychiatric disorder in NHS trust staff: occupational and gender differences. Br J Psychiatry. (1997) 171:519–23. doi: 10.1192/bjp.171.6.519

50. Olaya B, Perez-Moreno M, Bueno-Notivol J, Gracia-Garcia P, Lasheras I, Santabarbara J. Prevalence of depression among healthcare workers during the covid-19 outbreak: a systematic review and meta-analysis. J Clin Med. (2021) 10:3406. doi: 10.3390/jcm10153406

51. Bahamdan AS. Review of the psychological impact of COVID-19 pandemic on healthcare workers in Saudi Arabia. Risk Manag Healthc Policy. (2021) 14:4105–11. doi: 10.2147/RMHP.S324938

52. Norhayati MN, Che Yusof R, Azman MY. Prevalence of psychological impacts on healthcare providers during COVID-19 pandemic in Asia. Int J Environ Res Public Health. (2021) 18:9157. doi: 10.3390/ijerph18179157

53. Ching SM, Ng KY, Lee KW, Yee A, Lim PY, Ranita H, et al. Psychological distress among healthcare providers during COVID19 in Asia: systematic review and meta-analysis. PLoS ONE. (2021) 16:e0257983. doi: 10.1371/journal.pone.025798

54. Muntean M, Colcear D, Briciu V, et al. Comparison of the psychological impact of COVID-19 on healthcare workers between 2022 and 2023 in a Romanian COVID-19 Hub Hospital. COVID. 2024;4:1072–86

55. Pan L, Xu Q, Kuang X, et al. Prevalence and factors associated with posttraumatic stress disorder in healthcare workers exposed to COVID-19 in Wuhan, China: a cross-sectional survey. BMC Psychiatry. 2021;21(1):572.

56. Weathers FW, Litz BT, Herman DS, Huska JA, Keane TM. The PTSD Checklist (PCL): reliability, validity, and diagnostic utility. Paper presented at: Annual Convention of the International Society for Traumatic Stress Studies; 1993; San Antonio, TX.

57. Moodliar R, Russo J, Bedard-Gilligan M, et al. A pragmatic approach to psychometric comparisons between the DSM-IV and DSM-5 posttraumatic stress disorder (PTSD) checklists in acutely injured trauma patients. Psychiatry. 2020;83(4):390–401. doi: 10.1080/00332747.2020.1762396.

58. Moshier SJ, Lee DJ, Dovin MJ, Gauthier G, Zax A, Rosen RC, et al. An Empirical Crosswalk for the PTSD Checklist: Translating DSM-IV to DSM-5 Using a Veteran Sample. J Trauma Stress. 2019;32(5):799–805. doi: 10.1002/jts.22438

59. Becker MH. The Health Belief Model and sick role behavior. Health Educ Monogr. 1974;2(4):409–419. doi:10.1177/109019817400200407.

60. Hair JF Jr, Hult GTM, Ringle CM, Sarstedt M, Danks NP, Ray S. Partial least squares structural equation modeling (PLS-SEM) using R: a workbook. Cham: Springer; 2021. doi:10.1007/978-3-030-80519-7.. 62

61. Hair JF, Hult GTM, Ringle CM, Sarstedt M. *A Primer on Partial Least Squares Structural Equation Modeling (PLS-SEM)*. 2nd ed. Thousand Oaks, CA: Sage Publications; 2017.

62. Fornell C, Larcker DF. Evaluating structural equation models with unobservable variables and measurement error. J Mark Res. 1981;18(1):39–50.

63. Henseler J, Ringle CM, Sarstedt M. A new criterion for assessing discriminant validity in variance-based structural equation modeling. J Acad Mark Sci. 2015;43(1):115–135. doi:10.1007/s11747-014-0403-8.

64. Kock N. Common method bias in PLS-SEM: A full collinearity assessment approach. Int J E-Collab. 2015;11(4):1–10. doi:10.4018/ijec.2015100101

65. Carpenter CJ. A meta-analysis of the effectiveness of health belief model variables in predicting behavior. Health Commun. 2010;25(8):661–9.

